# Characteristics of individuals with cerebral palsy across the United States

**DOI:** 10.64898/2026.04.14.26350870

**Authors:** Bhooma R. Aravamuthan, Amy F. Bailes, Micah Baird, Kristie Bjornson, Ira Bowen, Angeline Bowman, Elizabeth R. Boyer, Rose Gelineau-Morel, Laurie Glader, Paul H Gross, Stacey Hall, Edward A. Hurvitz, Michael Kruer, Thomas Larrew, Neena Marupudi, Patrick G. McPhee, Stephen A Nichols, Garey Noritz, Joyce Oleszek, Justin Ramsey, Jeffrey Raskin, Heather Riordan, Brandon Rocque, Manish Shah, Benjamin J Shore, M. Wade Shrader, David Spence, Charles B. Stevenson, Richard Stevenson, Sruthi P. Thomas, Joyce P Trost, Stephen R. Wisniewski, the Cerebral Palsy Research Network

## Abstract

**Objective:** Cerebral palsy (CP) affects approximately 1 million Americans and 18 million individuals worldwide, yet contemporary US epidemiologic data remains limited. We aimed to use Cerebral Palsy Research Network (CPRN) clinical registry to describe demographics and clinical characteristics of individuals with CP across the US and determine associations with gross motor function and genetic etiology.

**Methods:** Registry subjects were included if they had clinician-confirmed CP and prospectively entered data for Gross Motor Function Classification System (GMFCS) Level, gestational age, genetic etiology, CP distribution, and tone/movement types. Logistic regression was used to determine which of these variables plus race, sex, ethnicity, and age were associated with GMFCS level and genetic etiology.

**Results:** A total of 9,756 children and adults with CP from 22 CPRN sites met inclusion criteria. Participants were predominantly White (73.0%), male (57.3%), non-Hispanic (87.8%), and younger than 18 years (73.7%). Most were classified as GMFCS levels I–III (55.6%), born preterm (52.8%), had spasticity (83.8%), and had quadriplegia (41.9%); 12.2% were identified as having a genetic etiology. Tone/movement types, CP distribution, and gestational age were significantly associated with both GMFCS level and genetic etiology (p<0.001). Compared to White individuals, Black individuals were more likely to have greater gross motor impairment (p<0.001).

**Conclusion:** In this large US cohort, clinical and demographic factors, including race, were associated with gross motor function and genetic etiology in CP. These findings highlight persistent disparities and demonstrate the value of a national clinical registry for informing prognostication, quality improvement efforts, and targeted genetic testing strategies.

---

Cerebral palsy (CP) is the most common childhood-onset motor disability, affecting approximately 1 million people in the US and 18 million people worldwide.^1^ Data on how CP presents largely comes from outside of the US.^2^ Descriptions of CP in the US through the Centers for Disease Control and Prevention (CDC) have been invaluable for estimating CP prevalence. However, because these estimates largely rely on regional retrospective chart reviews, they cannot provide detailed reports of other important CP characteristics like Gross Motor Function System Classification Level (GMFCS) and the presence or absence of a genetic etiology.^3^ Analysis of prospective clinician-reported data on individuals with CP remains limited across the US and across clinical sites. The lack of these data prevents a detailed understanding of CP in the US, which limits our understanding of gaps in care and impedes representative recruitment for clinical trials.

Using data prospectively entered by clinicians into the Cerebral Palsy Research Network (CPRN) clinical registry, we report on GMFCS Levels, CP distribution, tone/movement types, presence or absence of a genetic etiology, and demographic characteristics of a large cohort of individuals with CP across the US. This large prospective data set provides a unique clinician-driven overview of how CP manifests in the US and may help guide future studies to advance CP care in this country.

## Methods

Institutional Review Board (IRB) approval was granted by Nationwide Children’s Hospital with subsidiary reliance agreements for some CPRN-affiliated sites while other sites obtained individual IRB approval (See Supplementary Methods for list of sites and IRB approval numbers). As all data transmitted to the CPRN Data Coordinating Center do not include identifiable information and are extracted from the medical record, waiver of written consent was granted by all IRBs.

### Reported CPRN clinical registry data elements

The CPRN is comprised of 34 US tertiary-care clinical sites focused on conducting high quality clinical research and quality improvement for CP, including via pooling their clinical care data in a centralized registry. All individuals in this registry must meet criteria for a CP diagnosis according to the treating clinician’s assessment.^4^ Required and high priority clinician-reported data elements were determined by the CPRN using the National Institute of Neurological Disorders and Stroke Common Data Elements for CP^5^ and via iterative discussions between site investigators regarding the feasibility and value of routine entry of these elements during clinical care.^6,7^ These clinician-reported elements are the GMFCS level (I-V, with higher GMFCS levels indicating greater gross motor functional impairment),^8^ gestational age (<37 weeks or ≥37 weeks), genetic etiology (presence or absence), CP distribution (monoplegia, hemiplegia, diplegia, triplegia, or quadriplegia), and presence of dystonia, spasticity, hypotonia, or other tone/movement types (ataxia, chorea, and athetosis). Data were abstracted from the registry on 8/25/2025 from all individuals across sites who had a clinician-reported GMFCS level and at least one other clinician-reported element. A single value for each element was abstracted per individual, updated as of their most recent clinical encounter, with all sites updating this data centrally in the CPRN registry every 6 months.

Our analysis relied on any identification of tone/movement type(s) and not on the determination of the predominant type, which has been shown to be inconsistent even amongst expert assessors.^9,10^ Race (Black, White, or other which includes American Indian or Alaskan, Asian, and Native Hawaiian), ethnicity (Hispanic/Latino or not Hispanic/Latino), sex, and age were abstracted from the electronic health records by a site staff member or via the PCORnet Common Data Model.^11^

### Statistical analyses

Statistical analyses were done using SPSS (IBM, Armonk, NY, version 29). Chi-square tests were used to compare demographics / CP characteristics between all individuals (those with and without missing data) and individuals with complete data sets (no missing data). Data from individuals with complete data sets were further analyzed to determine associations with GMFCS level and the presence of a genetic etiology. Multiple logistic regression was used to determine whether race, gender, ethnicity, age, genetic etiology, gestational age, and tone/movement types were independently associated with GMFCS level. An ordinal regression model was not used because the assumption of proportional odds was not satisfied (test of parallel lines, *p*<0.001). Binary logistic regression was used to determine whether race, gender, ethnicity, age, gestational age, and tone/movement types, and GMFCS level affected the odds of having a genetic etiology of CP. Chi-square statistics were used to determine the significance of logistic regression models and model terms (*p*<0.05). Adjusted odds ratios (ORs) and 95% CIs were calculated for each variable. Variance inflation factors (VIFs) less than 5 were interpreted as the absence of multicollinearity.^12^ ANOVA and t-tests, respectively, were used to determine whether the number of documented tone/movement types differed between GMFCS levels or with identification of a genetic etiology (*p*<0.05).

## Results

As of 8/25/2025, data for 9,756 individuals with CP were available in the CPRN registry from 22 tertiary-care clinical sites across 17 states, including all 4 US Census regions and divisions (Supplementary Table 1).^13^ Demographics and missing data rates are in Table 1. With missing data excluded, most individuals were White (73.0%), male (57.3%), non-Hispanic (87.8%), and <18 years old (73.7%). The single most common CP distribution was quadriplegia (41.9%). Almost all had spasticity (83.8%). There was a contributing genetic etiology for 12.2% and half were born preterm (52.4% at <37 weeks gestation).

**Table 1.**
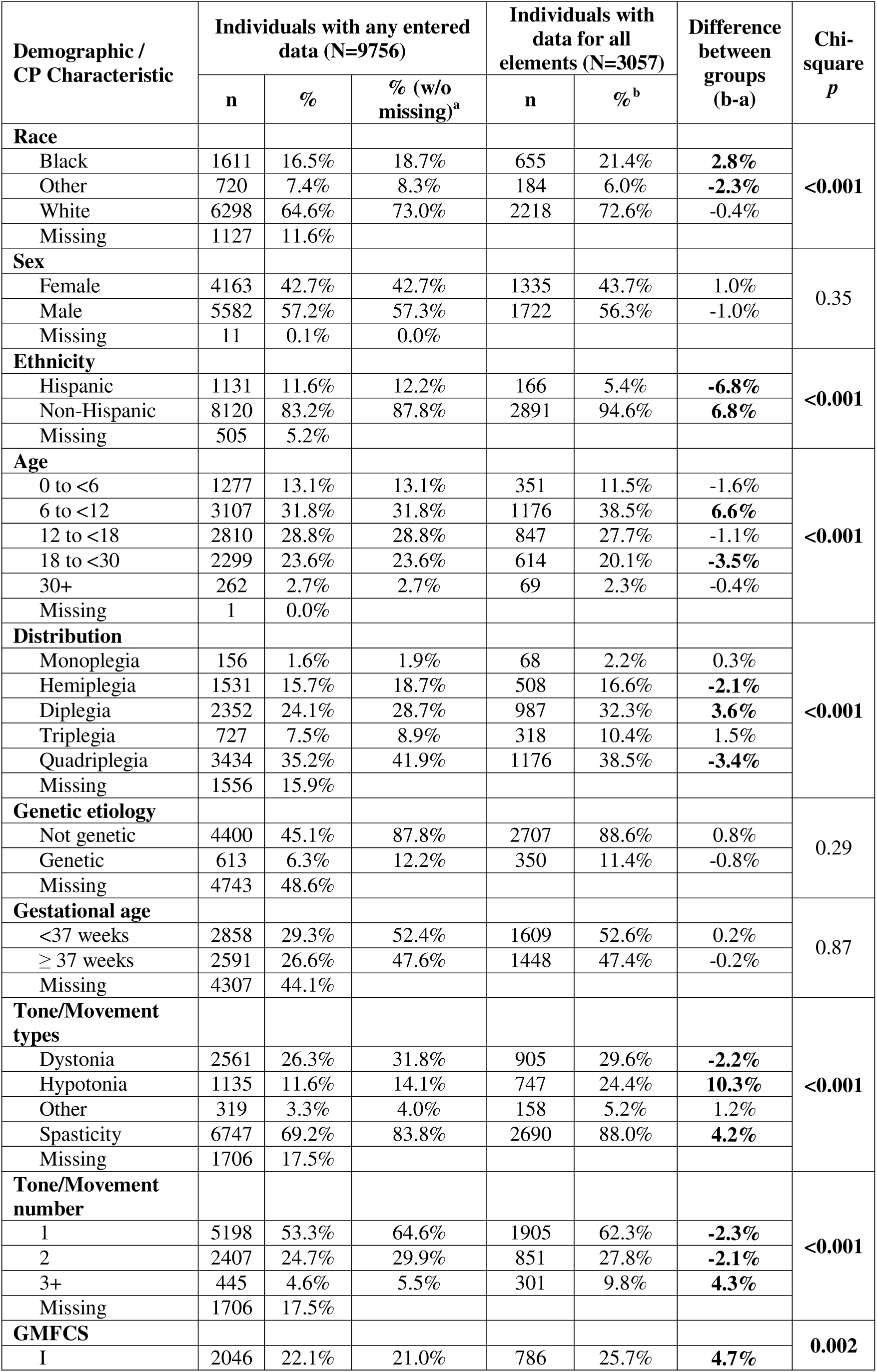
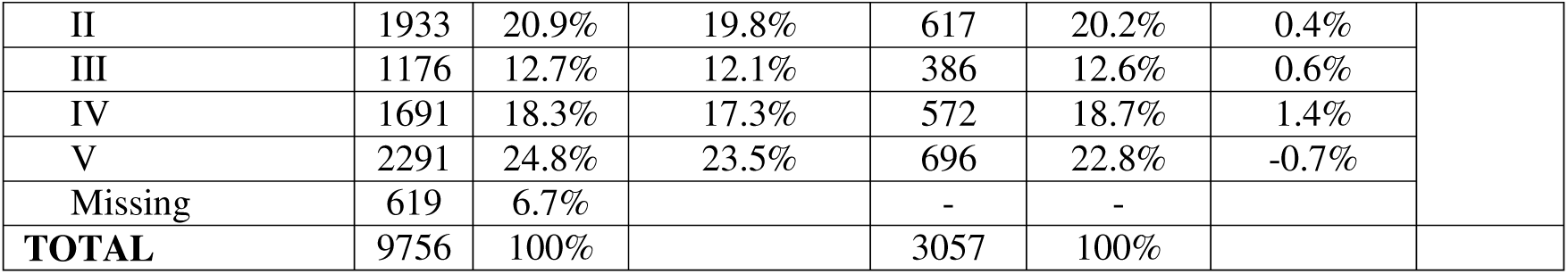
Demographics and CP Characteristics. Race (Black, White, or Other which includes American Indian or Alaskan, Asian, and Native Hawaiian), ethnicity (Hispanic/Latino or not Hispanic/Latino), sex, and age were abstracted from the electronic health records of these sites either by a dedicated staff member at each site or via the PCORnet Common Data Model. CP distribution (monoplegia, hemiplegia, diplegia, triplegia, or quadriplegia), presence of a contributing genetic etiology, gestational age (either less than 37 weeks or 37 weeks or more), tone/movement types present (dystonia, spasticity, hypotonia, or other movement types including ataxia, chorea, and athetosis), and Gross Motor Function Classification System level (GMFCS)^8^ were all prospectively entered by the treating clinician. Differences in proportions between individuals with data entered for all elements (n=3057)^b^ and individuals with any prospectively entered data (n=9756)^a^ are shown (differences greater than 2% in bold) and compared using a Chi-square test (*p*<0.05 in bold).

| Demographic /<br>CP Characteristic | Individuals with any entered<br>data (N=9756) |  |  | Individuals with<br>data for all<br>elements (N=3057) |  | Difference<br>between<br>groups<br>(b-a) | Chi-square<br>*p* |
| --- | --- | --- | --- | --- | --- | --- | --- |
|  | n | % | % (w/o<br>missing)<sup>a</sup> | n | %<sup>b</sup> |  |  |
| **Race** |  |  |  |  |  |  | <0.001 |
| Black | 1611 | 16.5% | 18.7% | 655 | 21.4% | **2.8%** |  |
| Other | 720 | 7.4% | 8.3% | 184 | 6.0% | **-2.3%** |  |
| White | 6298 | 64.6% | 73.0% | 2218 | 72.6% | -0.4% |  |
| Missing | 1127 | 11.6% |  |  |  |  |  |
| **Sex** |  |  |  |  |  |  | 0.35 |
| Female | 4163 | 42.7% | 42.7% | 1335 | 43.7% | 1.0% |  |
| Male | 5582 | 57.2% | 57.3% | 1722 | 56.3% | -1.0% |  |
| Missing | 11 | 0.1% | 0.0% |  |  |  |  |
| **Ethnicity** |  |  |  |  |  |  | <0.001 |
| Hispanic | 1131 | 11.6% | 12.2% | 166 | 5.4% | **-6.8%** |  |
| Non-Hispanic | 8120 | 83.2% | 87.8% | 2891 | 94.6% | **6.8%** |  |
| Missing | 505 | 5.2% |  |  |  |  |  |
| **Age** |  |  |  |  |  |  | <0.001 |
| 0 to <6 | 1277 | 13.1% | 13.1% | 351 | 11.5% | -1.6% |  |
| 6 to <12 | 3107 | 31.8% | 31.8% | 1176 | 38.5% | **6.6%** |  |
| 12 to <18 | 2810 | 28.8% | 28.8% | 847 | 27.7% | -1.1% |  |
| 18 to <30 | 2299 | 23.6% | 23.6% | 614 | 20.1% | **-3.5%** |  |
| 30+ | 262 | 2.7% | 2.7% | 69 | 2.3% | -0.4% |  |
| Missing | 1 | 0.0% |  |  |  |  |  |
| **Distribution** |  |  |  |  |  |  | <0.001 |
| Monoplegia | 156 | 1.6% | 1.9% | 68 | 2.2% | 0.3% |  |
| Hemiplegia | 1531 | 15.7% | 18.7% | 508 | 16.6% | **-2.1%** |  |
| Diplegia | 2352 | 24.1% | 28.7% | 987 | 32.3% | **3.6%** |  |
| Triplegia | 727 | 7.5% | 8.9% | 318 | 10.4% | 1.5% |  |
| Quadriplegia | 3434 | 35.2% | 41.9% | 1176 | 38.5% | **-3.4%** |  |
| Missing | 1556 | 15.9% |  |  |  |  |  |
| **Genetic etiology** |  |  |  |  |  |  | 0.29 |
| Not genetic | 4400 | 45.1% | 87.8% | 2707 | 88.6% | 0.8% |  |
| Genetic | 613 | 6.3% | 12.2% | 350 | 11.4% | -0.8% |  |
| Missing | 4743 | 48.6% |  |  |  |  |  |
| **Gestational age** |  |  |  |  |  |  | 0.87 |
| <37 weeks | 2858 | 29.3% | 52.4% | 1609 | 52.6% | 0.2% |  |
| ≥ 37 weeks | 2591 | 26.6% | 47.6% | 1448 | 47.4% | -0.2% |  |
| Missing | 4307 | 44.1% |  |  |  |  |  |
| **Tone/Movement**<br>**types** |  |  |  |  |  |  | <0.001 |
| Dystonia | 2561 | 26.3% | 31.8% | 905 | 29.6% | **-2.2%** |  |
| Hypotonia | 1135 | 11.6% | 14.1% | 747 | 24.4% | **10.3%** |  |
| Other | 319 | 3.3% | 4.0% | 158 | 5.2% | 1.2% |  |
| Spasticity | 6747 | 69.2% | 83.8% | 2690 | 88.0% | **4.2%** |  |
| Missing | 1706 | 17.5% |  |  |  |  |  |
| **Tone/Movement**<br>**number** |  |  |  |  |  |  | <0.001 |
| 1 | 5198 | 53.3% | 64.6% | 1905 | 62.3% | **-2.3%** |  |
| 2 | 2407 | 24.7% | 29.9% | 851 | 27.8% | **-2.1%** |  |
| 3+ | 445 | 4.6% | 5.5% | 301 | 9.8% | **4.3%** |  |
| Missing | 1706 | 17.5% |  |  |  |  |  |
| **GMFCS** |  |  |  |  |  |  | 0.002 |
| I | 2046 | 22.1% | 21.0% | 786 | 25.7% | **4.7%** |  |

**Table 1.** Demographics and CP Characteristics.
|  |  |  |  |  |  |  |
| --- | --- | --- | --- | --- | --- | --- |
| II | 1933 | 20.9% | 19.8% | 617 | 20.2% | 0.4% |
| III | 1176 | 12.7% | 12.1% | 386 | 12.6% | 0.6% |
| IV | 1691 | 18.3% | 17.3% | 572 | 18.7% | 1.4% |
| V | 2291 | 24.8% | 23.5% | 696 | 22.8% | -0.7% |
| Missing | 619 | 6.7% |  | - | - |  |
| **TOTAL** | 9756 | 100% |  | 3057 | 100% |  |

GMFCS followed a U-shaped distribution, with most individuals at levels I (21.0%) and V (23.5%) and the fewest at level III (12.1%). Of these 9,756 individuals, 3,057 had data entered for all assessed registry elements (race, gender, ethnicity, age, CP distribution, genetic etiology, gestational age, tone/movement types, and GMFCS). These 3,057 individuals had generally comparable demographics to the full population but did have a significantly higher proportion of individuals who were Black (21.4% vs. 18.7%), non-Hispanic (94.6% vs. 87.8%), 6-12 years old (38.5% vs. 31.8%), had diplegia (32.3% vs.

28.7%), had hypotonia (24.4% vs. 14.1%), or were at GMFCS Level I (25.7% vs. 22.1%) (Chi-square statistic 1373, *p*<0.001, Table 1). The age was significantly lower in those with a complete data set (13.3 years, 95% CI 13.0-13.5) compared to those with any data entered (13.9 years, 95% CI 13.7-14.0) (t-test *p*<0.001).

### Associations with GMFCS Level

A multiple logistic regression model incorporating race, gender, ethnicity, age, CP distribution, genetic etiology, gestational age, and tone/movement types demonstrated significant associations with GMFCS level (overall model Chi-square statistic 1922, p<0.001) with VIFs between 1-1.6 indicating lack of multi-collinearity (Table 2). Race, age, CP distribution, gestational age, and all tone/movement types were independently associated with GMFCS Level (Table 3). Compared to individuals at GMFCS level V (greatest gross motor impairment), individuals at level I (least gross motor impairment) were significantly less likely to be Black (OR 0.48, 95% CI 0.35-0.67, p<0.001), were younger (OR 0.93 with age increase of 1 year, 95% CI 0.91-0.95, p<0.001), were more likely to have any CP distribution other than quadriplegia (p<0.001), were less likely to be born at term (OR 0.51, 95% CI 0.38-0.67, p<0.001), and were less likely to have the tone/movement types of dystonia (OR 0.13, 95% CI 0.09-0.18, p<0.001), hypotonia (OR 0.61, 95% CI 0.40-0.93, p=0.02), or spasticity (OR 0.11, 95% CI 0.07-0.19) (Table 3).

**Table 2.** Demographics and CP Characteristics across Gross Motor Function Classification System (GMFCS) Levels.

| Demographic/CP Characteristic | Level I |  |  | Level II |  |  | Level III |  |  | Level IV |  |  | Level V |  |  | Missing GMFCS |  |
| --- | --- | --- | --- | --- | --- | --- | --- | --- | --- | --- | --- | --- | --- | --- | --- | --- | --- |
|  | n | % | % (w/o missing) | n | % | % (w/o missing) | n | % | % (w/o missing) | n | % | % (w/o missing) | n | % | % (w/o missing) | n | % |
| Race |  |  |  |  |  |  |  |  |  |  |  |  |  |  |  |  |  |
| Black | 273 | 13.3% | 14.9% | 299 | 15.5% | 17.4% | 178 | 15.1% | 17.1% | 328 | 19.4% | 21.5% | 461 | 20.1% | 23.2% | 72 | 11.6% |
| Other | 152 | 7.4% | 8.3% | 132 | 6.8% | 7.7% | 80 | 6.8% | 7.7% | 102 | 6.0% | 6.7% | 164 | 7.2% | 8.2% | 90 | 14.5% |
| White | 1413 | 69.1% | 76.9% | 1285 | 66.5% | 74.9% | 780 | 66.3% | 75.1% | 1095 | 64.8% | 71.8% | 1363 | 59.5% | 68.6% | 362 | 58.5% |
| Missing Race | 208 | 10.2% |  | 217 | 11.2% |  | 138 | 11.7% |  | 166 | 9.8% |  | 303 | 13.2% |  | 95 | 15.3% |
| Sex |  |  |  |  |  |  |  |  |  |  |  |  |  |  |  |  |  |
| Female | 917 | 44.8% | 44.8% | 840 | 43.5% | 43.5% | 477 | 40.6% | 40.6% | 704 | 41.6% | 41.7% | 953 | 41.6% | 41.6% | 272 | 43.9% |
| Male | 1128 | 55.1% | 55.2% | 1090 | 56.4% | 56.5% | 698 | 59.4% | 59.4% | 983 | 58.1% | 58.3% | 1336 | 58.3% | 58.4% | 347 | 56.1% |
| Missing Sex | 1 | 0.0% |  | 3 | 0.2% |  | 1 | 0.1% |  | 4 | 0.2% |  | 2 | 0.1% |  | 0 | 0.0% |
| Ethnicity |  |  |  |  |  |  |  |  |  |  |  |  |  |  |  |  |  |
| Hispanic | 207 | 10.1% | 10.4% | 215 | 11.1% | 11.6% | 123 | 10.5% | 10.9% | 173 | 10.2% | 10.7% | 328 | 14.3% | 15.1% | 85 | 13.7% |
| Non-Hispanic | 1777 | 86.9% | 89.6% | 1642 | 84.9% | 88.4% | 1006 | 85.5% | 89.1% | 1438 | 85.0% | 89.3% | 1846 | 80.6% | 84.9% | 411 | 66.4% |
| Missing Ethnicity | 62 | 3.0% |  | 76 | 3.9% |  | 47 | 4.0% |  | 80 | 4.7% |  | 117 | 5.1% |  | 123 | 19.9% |
| Age (years) |  |  |  |  |  |  |  |  |  |  |  |  |  |  |  |  |  |
| 0 to <6 | 190 | 9.3% | 9.3% | 171 | 8.8% | 8.8% | 129 | 11.0% | 11.0% | 147 | 8.7% | 8.7% | 212 | 9.3% | 9.3% | 428 | 69.1% |
| 6 to <12 | 767 | 37.5% | 37.5% | 644 | 33.3% | 33.3% | 442 | 37.6% | 37.6% | 532 | 31.5% | 31.5% | 644 | 28.1% | 28.1% | 78 | 12.6% |
| 12 to <18 | 646 | 31.6% | 31.6% | 611 | 31.6% | 31.6% | 322 | 27.4% | 27.4% | 478 | 28.3% | 28.3% | 703 | 30.7% | 30.7% | 50 | 8.1% |
| 18 to <30 | 404 | 19.7% | 19.7% | 457 | 23.6% | 23.6% | 240 | 20.4% | 20.4% | 474 | 28.0% | 28.0% | 669 | 29.2% | 29.2% | 55 | 8.9% |
| 30+ | 39 | 1.9% | 1.9% | 50 | 2.6% | 2.6% | 43 | 3.7% | 3.7% | 60 | 3.5% | 3.5% | 62 | 2.7% | 2.7% | 8 | 1.3% |
| Missing Age | 0 | 0.0% |  | 0 | 0.0% |  | 0 | 0.0% |  | 0 | 0.0% |  | 1 | 0.0% |  | 0 |  |
| CP Distribution |  |  |  |  |  |  |  |  |  |  |  |  |  |  |  |  |  |
| Monoplegia | 99 | 4.8% | 5.6% | 32 | 1.7% | 1.9% | 5 | 0.4% | 0.5% | 8 | 0.5% | 0.5% | 3 | 0.1% | 0.2% | 9 | 1.5% |
| Hemiplegia | 795 | 38.9% | 45.2% | 552 | 28.6% | 32.9% | 62 | 5.3% | 6.0% | 49 | 2.9% | 3.3% | 14 | 0.6% | 0.7% | 59 | 9.5% |
| Diplegia | 618 | 30.2% | 35.1% | 702 | 36.3% | 41.9% | 517 | 44.0% | 50.0% | 297 | 17.6% | 19.9% | 160 | 7.0% | 8.1% | 58 | 9.4% |
| Triplegia | 128 | 6.3% | 7.3% | 181 | 9.4% | 10.8% | 164 | 13.9% | 15.8% | 189 | 11.2% | 12.7% | 49 | 2.1% | 2.5% | 16 | 2.6% |
| Quadriplegia | 120 | 5.9% | 6.8% | 210 | 10.9% | 12.5% | 287 | 24.4% | 27.7% | 946 | 55.9% | 63.5% | 1758 | 76.7% | 88.6% | 113 | 18.3% |
| Missing Dist. | 286 | 14.0% |  | 256 | 13.2% |  | 141 | 12.0% |  | 202 | 11.9% |  | 307 | 13.4% |  | 364 | 58.8% |
| Genetic etiology |  |  |  |  |  |  |  |  |  |  |  |  |  |  |  |  |  |
| Not genetic | 1157 | 56.5% | 94.1% | 934 | 48.3% | 86.5% | 521 | 44.3% | 86.4% | 743 | 43.9% | 84.8% | 925 | 40.4% | 84.9% | 120 | 19.4% |
| Genetic | 73 | 3.6% | 5.9% | 146 | 7.6% | 13.5% | 82 | 7.0% | 13.6% | 133 | 7.9% | 15.2% | 164 | 7.2% | 15.1% | 15 | 2.4% |
| Missing etiology | 816 | 39.9% |  | 853 | 44.1% |  | 573 | 48.7% |  | 815 | 48.2% |  | 1202 | 52.5% |  | 484 | 78.2% |
| Gestational age |  |  |  |  |  |  |  |  |  |  |  |  |  |  |  |  |  |
| <37 weeks | 678 | 33.1% | 53.6% | 559 | 28.9% | 52.0% | 419 | 35.6% | 63.4% | 532 | 31.5% | 57.3% | 486 | 21.2% | 40.8% | 184 | 29.7% |
| ≥ 37 weeks | 588 | 28.7% | 46.4% | 515 | 26.6% | 48.0% | 242 | 20.6% | 36.6% | 396 | 23.4% | 42.7% | 706 | 30.8% | 59.2% | 144 | 23.3% |
| Missing gest. age | 780 | 38.1% |  | 859 | 44.4% |  | 515 | 43.8% |  | 763 | 45.1% |  | 1099 | 48.0% |  | 291 | 47.0% |
| Tone/Movement type |  |  |  |  |  |  |  |  |  |  |  |  |  |  |  |  |  |
| Dystonia | 244 | 11.9% | 14.8% | 418 | 21.6% | 28.6% | 308 | 26.2% | 31.4% | 599 | 35.4% | 42.4% | 909 | 39.7% | 46.7% | 83 | 13.4% |
| Hypotonia | 171 | 8.4% | 10.4% | 199 | 10.3% | 13.6% | 158 | 13.4% | 16.1% | 241 | 14.3% | 17.0% | 331 | 14.4% | 17.0% | 35 | 5.7% |
| Other | 52 | 2.5% | 3.1% | 79 | 4.1% | 5.4% | 60 | 5.1% | 6.1% | 60 | 3.5% | 4.2% | 61 | 2.7% | 3.1% | 7 | 1.1% |
| Spasticity | 1402 | 68.5% | 84.9% | 1243 | 64.3% | 85.1% | 861 | 73.2% | 87.8% | 1246 | 73.7% | 88.1% | 1738 | 75.9% | 89.3% | 257 | 41.5% |
| Missing tone | 395 | 19.3% |  | 472 | 24.4% |  | 195 | 16.6% |  | 277 | 16.4% |  | 344 | 15.0% |  | 23 | 3.7% |
| **Tone/Movement number** |  |  |  |  |  |  |  |  |  |  |  |  |  |  |  |  |  |
| 1 | 1366 | 66.8% | 82.7% | 981 | 50.8% | 67.1% | 610 | 51.9% | 62.2% | 751 | 44.4% | 53.1% | 983 | 42.9% | 50.5% | 507 | 81.9% |
| 2 | 266 | 13.0% | 16.1% | 427 | 22.1% | 29.2% | 316 | 26.9% | 32.2% | 555 | 32.8% | 39.3% | 762 | 33.3% | 39.1% | 81 | 13.1% |
| 3+ | 19 | 0.9% | 1.2% | 53 | 2.7% | 3.6% | 55 | 4.7% | 5.6% | 108 | 6.4% | 7.6% | 202 | 8.8% | 10.4% | 8 | 1.3% |
| Missing | 395 | 19.3% |  | 472 | 24.4% |  | 195 | 16.6% |  | 277 | 16.4% |  | 344 | 15.0% |  | 23 | 3.7% |
| **TOTAL** | 2046 | 100% |  | 1933 | 100% |  | 1176 | 100% |  | 1691 | 100% |  | 2291 | 100% |  | 619 | 100% |

**Table 3.** Odds ratios for demographics and CP characteristics across Gross Motor Classification System (GMFCS) Levels I-IV, as compared to Level V. Multiple logistic regression odds ratios with 95% confidence intervals (CI) and corresponding Chi-square *p-*values are shown (*p*<0.05 in bold). Reference categories are indicated for categorical variables of race (Black, Other including American Indian or Alaskan, Asian, and Native Hawaiian, and White, ^a^Overall *p*-value 0.001) and CP distribution (monoplegia, hemiplegia, diplegia, triplegia, quadriplegia, ^b^Overall *p*-value <0.001). E.g. a Black person is 0.48 (95% CI 0.35-0.67) times as likely as a White person to be at Level I vs. Level V. Sex (male, female), ethnicity (Hispanic, Non-Hispanic), genetic etiology (present, absent), and gestational age (term ≥ 37 weeks, preterm <37 weeks) were all analyzed as binary variables. E.g. a person with gestational age ≥37 weeks is 0.51 (95% CI 0.38-0.67) times as likely as a person with gestational age <37 weeks to be at Level I vs. Level V. Age was analyzed as a continuous variable. E.g. for every 1 year increase in age, a person is 0.93 (95% CI 0.91-0.95) times as likely to be at Level I vs. Level V.

| Demographic/ CP Characteristic | OR for each GMFCS Level (Reference: Level V) |  |  |  |  |  |  |  |  |  |  |  |  |  |  |  |
| --- | --- | --- | --- | --- | --- | --- | --- | --- | --- | --- | --- | --- | --- | --- | --- | --- |
|  | Level I |  |  |  | Level II |  |  |  | Level III |  |  |  | Level IV |  |  |  |
|  | OR | 95% CI for OR |  | *p* | OR | 95% CI for OR |  | *p* | OR | 95% CI for OR |  | *p* | OR | 95% CI for OR |  | *p* |
|  |  | Lower | Upper |  |  | Lower | Upper |  |  | Lower | Upper |  |  | Lower | Upper |  |
| Race: Black (Ref: White)<sup>a</sup> | 0.48 | 0.35 | 0.67 | **0.00** | 0.66 | 0.48 | 0.91 | **0.01** | 0.70 | 0.50 | 0.98 | **0.04** | 0.88 | 0.67 | 1.16 | 0.36 |
| Race: Other (Ref: White)<sup>a</sup> | 0.75 | 0.44 | 1.27 | 0.28 | 0.50 | 0.28 | 0.90 | **0.02** | 0.90 | 0.53 | 1.56 | 0.72 | 0.66 | 0.39 | 1.10 | 0.11 |
| Sex: Male | 0.87 | 0.67 | 1.14 | 0.31 | 0.84 | 0.64 | 1.09 | 0.19 | 1.00 | 0.76 | 1.32 | 0.98 | 1.00 | 0.79 | 1.26 | 0.97 |
| Ethnicity: Hispanic | 0.97 | 0.53 | 1.79 | 0.93 | 1.14 | 0.64 | 2.05 | 0.65 | 1.27 | 0.71 | 2.27 | 0.43 | 0.94 | 0.57 | 1.55 | 0.82 |
| Age (Increase by 1 year) | 0.93 | 0.91 | 0.95 | **0.00** | 0.97 | 0.95 | 0.99 | **0.00** | 0.95 | 0.93 | 0.97 | **0.00** | 0.99 | 0.98 | 1.01 | 0.28 |
| Monoplegia (Ref: Quadriplegia)<sup>b</sup> | 284.18 | 64.46 | 1252.95 | **0.00** | 78.65 | 17.23 | 358.93 | **0.00** | 9.88 | 1.60 | 61.08 | **0.01** | 3.68 | 0.66 | 20.42 | 0.14 |
| Hemiplegia (Ref: Quadriplegia)<sup>b</sup> | 1666.45 | 505.12 | 5497.87 | **0.00** | 771.44 | 236.71 | 2514.12 | **0.00** | 47.68 | 13.83 | 164.37 | **0.00** | 11.36 | 3.26 | 39.51 | **0.00** |
| Diplegia (Ref: Quadriplegia)<sup>b</sup> | 29.02 | 19.73 | 42.70 | **0.00** | 17.61 | 12.30 | 25.22 | **0.00** | 7.40 | 5.27 | 10.39 | **0.00** | 1.91 | 1.41 | 2.58 | **0.00** |
| Triplegia (Ref: Quadriplegia)<sup>b</sup> | 46.70 | 26.18 | 83.33 | **0.00** | 34.48 | 19.82 | 60.00 | **0.00** | 13.32 | 7.69 | 23.06 | **0.00** | 6.20 | 3.76 | 10.22 | **0.00** |
| Genetic etiology | 0.62 | 0.39 | 1.00 | 0.05 | 1.10 | 0.73 | 1.68 | 0.64 | 1.25 | 0.81 | 1.93 | 0.31 | 1.04 | 0.72 | 1.49 | 0.85 |
| Gestational age: ≥37 weeks | 0.51 | 0.38 | 0.67 | **0.00** | 0.61 | 0.47 | 0.81 | **0.00** | 0.42 | 0.31 | 0.55 | **0.00** | 0.53 | 0.42 | 0.67 | **0.00** |
| Tone/Movement: Dystonia | 0.13 | 0.09 | 0.18 | **0.00** | 0.38 | 0.29 | 0.52 | **0.00** | 0.54 | 0.40 | 0.72 | **0.00** | 0.68 | 0.53 | 0.87 | **0.00** |
| Tone/Movement: Hypotonia | 0.61 | 0.40 | 0.93 | **0.02** | 1.11 | 0.76 | 1.61 | 0.59 | 0.81 | 0.56 | 1.18 | 0.27 | 0.96 | 0.71 | 1.29 | 0.78 |
| Tone/Movement: Other | 1.64 | 0.83 | 3.23 | 0.15 | 2.66 | 1.47 | 4.84 | **0.00** | 2.73 | 1.50 | 4.96 | **0.00** | 1.81 | 1.04 | 3.15 | **0.04** |
| Tone/Movement: Spasticity | 0.11 | 0.07 | 0.19 | **0.00** | 0.18 | 0.11 | 0.29 | **0.00** | 0.33 | 0.20 | 0.55 | **0.00** | 0.63 | 0.41 | 0.97 | **0.04** |

These associations trended with GMFCS level. That is, with increasing GMFCS level, the proportion of individuals who are Black, older, had quadriplegia, or had dystonia, hypotonia, or spasticity increased (Table 2, Figure 1). The number of tone/movement types documented in the same individual also significantly increased with increasing GMFCS level (ANOVA F-statistic 100, p<0.001) (Table 2).

**Figure 1.**
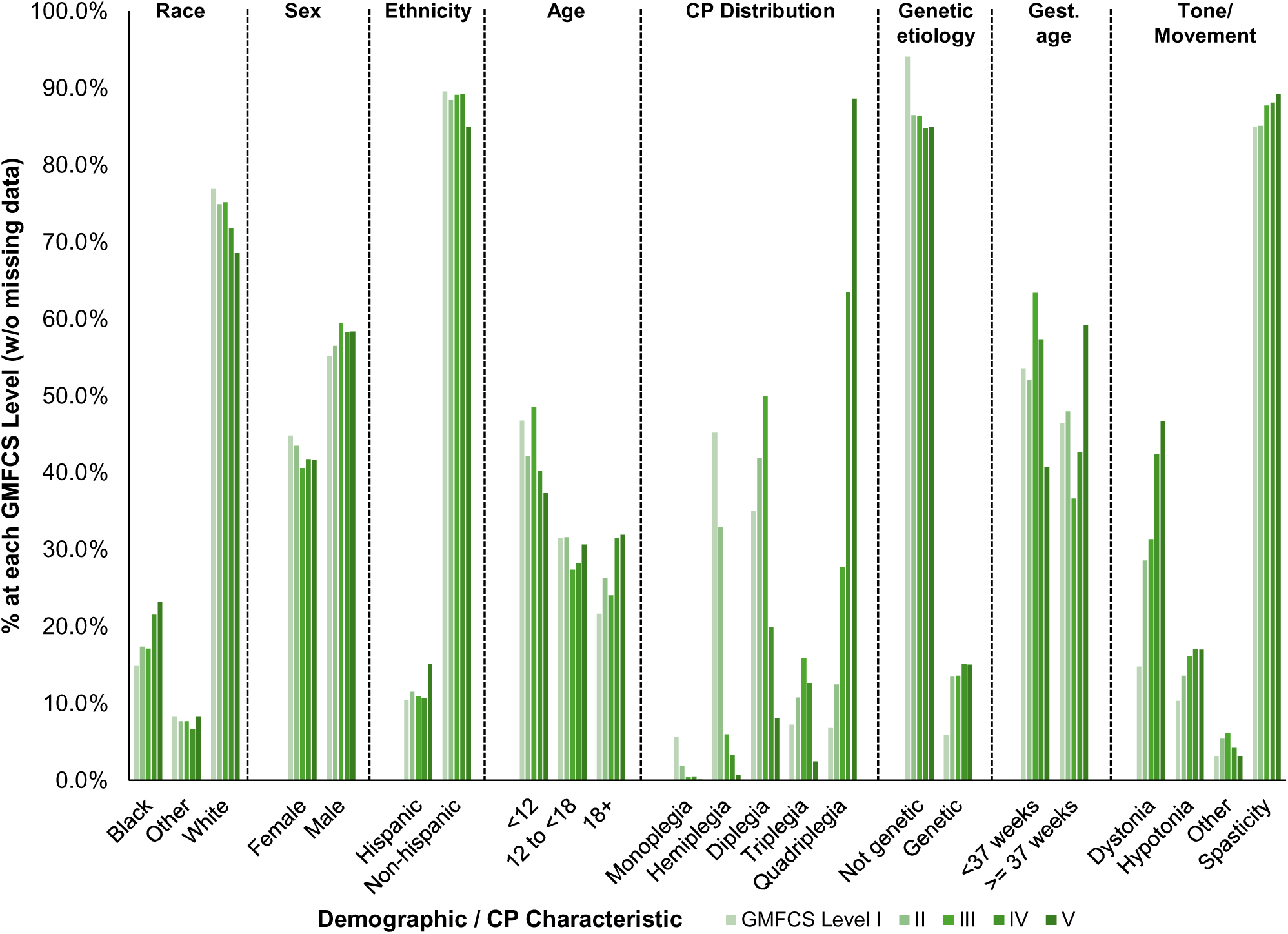
Demographics and CP Characteristics across Gross Motor Function Classification System (GMFCS) Levels.

Notably, when comparing GMFCS levels I and V, the differences in proportions of individuals with dystonia (14.8% at level I vs. 46.7% at level V) and hypotonia (10.4% vs. 17.0%) were much larger than the difference in the proportion of individuals with spasticity (84.9% vs. 89.3%) (Table 2). Dystonia almost always occurred in combination with other tone/movement types regardless of GMFCS level (90-95% of those with dystonia, Table 4). However, hypotonia (62%) and spasticity (81%) were typically described in isolation at GMFCS level I but typically described in combination with other tone/movement types at GMFCS level V (89% of those with hypotonia and 53% of those with spasticity) (Table 4).

**Table 4.** Percentage of individuals with each tone/movement type present in isolation (only) or in combination (mixed) stratified by GMFCS level.

| GMFCS | Dystonia |  |  |  | Hypotonia |  |  |  | Other (Ataxia, Chorea, Athetosis) |  |  |  | Spasticity |  |  |  |
| --- | --- | --- | --- | --- | --- | --- | --- | --- | --- | --- | --- | --- | --- | --- | --- | --- |
|  | Only | % | Mixed | % | Only | % | Mixed | % | Only | % | Mixed | % | Only | % | Mixed | % |
| I | 25 | 10% | 219 | 90% | 106 | 62% | 65 | 38% | 15 | 29% | 37 | 71% | 1133 | 81% | 269 | 19% |
| II | 33 | 8% | 385 | 92% | 78 | 39% | 121 | 61% | 16 | 20% | 63 | 80% | 796 | 64% | 447 | 36% |
| III | 15 | 5% | 293 | 95% | 41 | 26% | 117 | 74% | 10 | 17% | 50 | 83% | 522 | 61% | 339 | 39% |
| IV | 36 | 6% | 563 | 94% | 47 | 20% | 194 | 80% | 7 | 12% | 53 | 88% | 620 | 50% | 626 | 50% |
| V | 43 | 5% | 866 | 95% | 35 | 11% | 296 | 89% | 4 | 7% | 57 | 93% | 819 | 47% | 919 | 53% |

### Associations with genetic etiology

A binary logistic regression model incorporating race, gender, ethnicity, age, CP distribution, gestational age, tone/movement types, and GMFCS level demonstrated significant associations with the presence of a genetic etiology (overall model Chi-square statistic 462, p<0.001) with VIFs between 1-1.8 indicating lack of multi-collinearity (Table 5). Individuals with a genetic etiology for CP, compared to those without, were significantly less likely to have triplegia (OR 0.48, 95% CI 0.26-0.88, p=0.02) and less likely to have spasticity (OR 0.68, 95% CI 0.48-0.96, p=0.03). They were significantly more likely to be born at term (OR 4.68, 95% CI 3.52-6.24, p<0.001) and more likely to have hypotonia (OR 4.39, 95% CI 3.25-5.94, p<0.001) and other tone/movement types (ataxia, chorea, and athetosis, OR 2.16, 95% CI 1.40-3.33, p<0.001) (Table 6, Figure 2).

**Figure 2.**
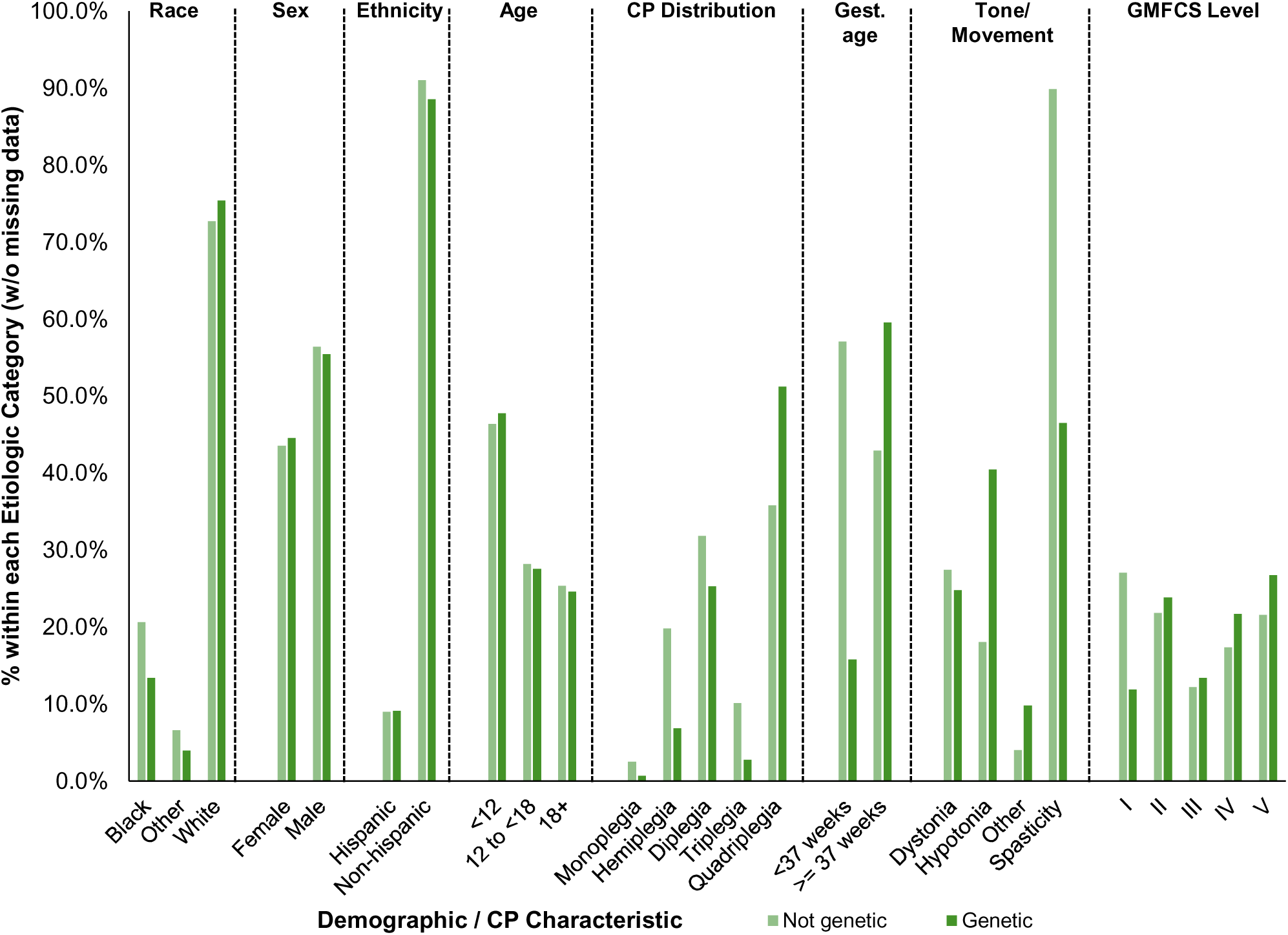
Demographics and CP characteristics across people with genetic and non-genetic etiologies. GMFCS – Gross Motor Function Classification System.

**Table 5.**
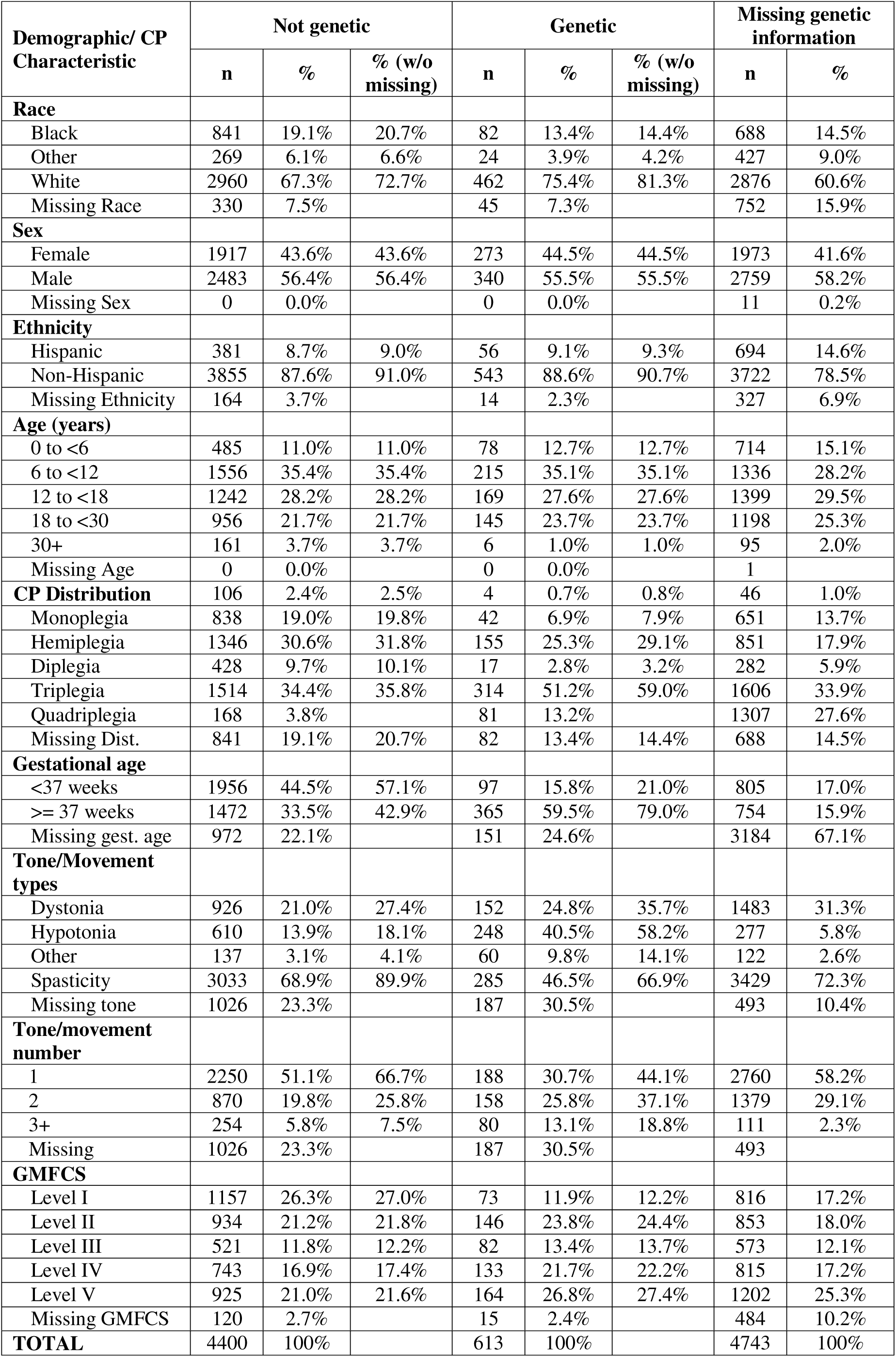
Demographics and CP characteristics in individuals with genetic and non-genetic etiologies.

| Demographic/ CP Characteristic | Not genetic |  |  | Genetic |  |  | Missing genetic information |  |
| --- | --- | --- | --- | --- | --- | --- | --- | --- |
|  | n | % | % (w/o missing) | n | % | % (w/o missing) | n | % |
| **Race** |  |  |  |  |  |  |  |  |
| Black | 841 | 19.1% | 20.7% | 82 | 13.4% | 14.4% | 688 | 14.5% |
| Other | 269 | 6.1% | 6.6% | 24 | 3.9% | 4.2% | 427 | 9.0% |
| White | 2960 | 67.3% | 72.7% | 462 | 75.4% | 81.3% | 2876 | 60.6% |
| Missing Race | 330 | 7.5% |  | 45 | 7.3% |  | 752 | 15.9% |
| **Sex** |  |  |  |  |  |  |  |  |
| Female | 1917 | 43.6% | 43.6% | 273 | 44.5% | 44.5% | 1973 | 41.6% |
| Male | 2483 | 56.4% | 56.4% | 340 | 55.5% | 55.5% | 2759 | 58.2% |
| Missing Sex | 0 | 0.0% |  | 0 | 0.0% |  | 11 | 0.2% |
| **Ethnicity** |  |  |  |  |  |  |  |  |
| Hispanic | 381 | 8.7% | 9.0% | 56 | 9.1% | 9.3% | 694 | 14.6% |
| Non-Hispanic | 3855 | 87.6% | 91.0% | 543 | 88.6% | 90.7% | 3722 | 78.5% |
| Missing Ethnicity | 164 | 3.7% |  | 14 | 2.3% |  | 327 | 6.9% |
| **Age (years)** |  |  |  |  |  |  |  |  |
| 0 to <6 | 485 | 11.0% | 11.0% | 78 | 12.7% | 12.7% | 714 | 15.1% |
| 6 to <12 | 1556 | 35.4% | 35.4% | 215 | 35.1% | 35.1% | 1336 | 28.2% |
| 12 to <18 | 1242 | 28.2% | 28.2% | 169 | 27.6% | 27.6% | 1399 | 29.5% |
| 18 to <30 | 956 | 21.7% | 21.7% | 145 | 23.7% | 23.7% | 1198 | 25.3% |
| 30+ | 161 | 3.7% | 3.7% | 6 | 1.0% | 1.0% | 95 | 2.0% |
| Missing Age | 0 | 0.0% |  | 0 | 0.0% |  | 1 |  |
| **CP Distribution** | 106 | 2.4% | 2.5% | 4 | 0.7% | 0.8% | 46 | 1.0% |
| Monoplegia | 838 | 19.0% | 19.8% | 42 | 6.9% | 7.9% | 651 | 13.7% |
| Hemiplegia | 1346 | 30.6% | 31.8% | 155 | 25.3% | 29.1% | 851 | 17.9% |
| Diplegia | 428 | 9.7% | 10.1% | 17 | 2.8% | 3.2% | 282 | 5.9% |
| Triplegia | 1514 | 34.4% | 35.8% | 314 | 51.2% | 59.0% | 1606 | 33.9% |
| Quadriplegia | 168 | 3.8% |  | 81 | 13.2% |  | 1307 | 27.6% |
| Missing Dist. | 841 | 19.1% | 20.7% | 82 | 13.4% | 14.4% | 688 | 14.5% |
| **Gestational age** |  |  |  |  |  |  |  |  |
| <37 weeks | 1956 | 44.5% | 57.1% | 97 | 15.8% | 21.0% | 805 | 17.0% |
| >= 37 weeks | 1472 | 33.5% | 42.9% | 365 | 59.5% | 79.0% | 754 | 15.9% |
| Missing gest. age | 972 | 22.1% |  | 151 | 24.6% |  | 3184 | 67.1% |
| **Tone/Movement types** |  |  |  |  |  |  |  |  |
| Dystonia | 926 | 21.0% | 27.4% | 152 | 24.8% | 35.7% | 1483 | 31.3% |
| Hypotonia | 610 | 13.9% | 18.1% | 248 | 40.5% | 58.2% | 277 | 5.8% |
| Other | 137 | 3.1% | 4.1% | 60 | 9.8% | 14.1% | 122 | 2.6% |
| Spasticity | 3033 | 68.9% | 89.9% | 285 | 46.5% | 66.9% | 3429 | 72.3% |
| Missing tone | 1026 | 23.3% |  | 187 | 30.5% |  | 493 | 10.4% |
| **Tone/movement number** |  |  |  |  |  |  |  |  |
| 1 | 2250 | 51.1% | 66.7% | 188 | 30.7% | 44.1% | 2760 | 58.2% |
| 2 | 870 | 19.8% | 25.8% | 158 | 25.8% | 37.1% | 1379 | 29.1% |
| 3+ | 254 | 5.8% | 7.5% | 80 | 13.1% | 18.8% | 111 | 2.3% |
| Missing | 1026 | 23.3% |  | 187 | 30.5% |  | 493 |  |
| **GMFCS** |  |  |  |  |  |  |  |  |
| Level I | 1157 | 26.3% | 27.0% | 73 | 11.9% | 12.2% | 816 | 17.2% |
| Level II | 934 | 21.2% | 21.8% | 146 | 23.8% | 24.4% | 853 | 18.0% |
| Level III | 521 | 11.8% | 12.2% | 82 | 13.4% | 13.7% | 573 | 12.1% |
| Level IV | 743 | 16.9% | 17.4% | 133 | 21.7% | 22.2% | 815 | 17.2% |
| Level V | 925 | 21.0% | 21.6% | 164 | 26.8% | 27.4% | 1202 | 25.3% |
| Missing GMFCS | 120 | 2.7% |  | 15 | 2.4% |  | 484 | 10.2% |
| **TOTAL** | 4400 | 100% |  | 613 | 100% |  | 4743 | 100% |

**Table 6.** Odds ratios for demographics and CP characteristics for people with genetic etiologies, compared to those with only non-genetic etiologies. Binary logistic regression odds ratios with 95% confidence intervals (CI) and corresponding Chi-square *p-*values are shown (*p*<0.05 in bold). Reference categories are indicated for categorical variables of race (Black, Other including American Indian or Alaskan, Asian, and Native Hawaiian, and White, ^a^Overall *p*-value=0.05), CP distribution (monoplegia, hemiplegia, diplegia, triplegia, quadriplegia, ^b^Overall *p*-value=0.06), and GMFCS Level (I-V, ^c^Overall *p*-value=0.03), reference categories are indicated. E.g. A person in the Other race category is 0.54 (95% CI 0.29-1.01) as likely as a White person to have a genetic CP etiology, though this relationship did not reach significance (*p*=0.05). Sex (male, female), ethnicity (Hispanic, Non-Hispanic), and gestational age (term ≥ 37 weeks, preterm <37 weeks) were all analyzed as binary variables. E.g. a person with gestational age ≥37 weeks is 4.68 (95% CI 3.52-6.24) times as likely as a person with gestational age <37 weeks to have a genetic etiology. Age was analyzed as a continuous variable. E.g. for every 1 year increase in age, a person is 1.01 (95% CI 0.99-1.03) times as likely to have a genetic etiology, though this relationship did not reach significance (*p*=0.51).

| Demographic/CP Characteristic | OR for Genetic etiology |  |  |  |
| --- | --- | --- | --- | --- |
|  | OR | 95% CI of OR |  | *P* |
|  |  | Lower | Upper |  |
| **Race: Black (Ref: White)<sup>a</sup>** | 0.76 | 0.54 | 1.06 | 0.10 |
| **Race: Other (Ref: White)<sup>a</sup>** | 0.54 | 0.29 | 1.01 | 0.05 |
| **Sex: Male** | 1.08 | 0.84 | 1.39 | 0.54 |
| **Ethnicity: Hispanic** | 0.62 | 0.33 | 1.17 | 0.14 |
| **Age (Increase by 1 year)** | 1.01 | 0.99 | 1.03 | 0.51 |
| **Monoplegia (Ref: Quadriplegia)<sup>b</sup>** | 0.00 | 0.00 |  | 1.00 |
| **Hemiplegia (Ref: Quadriplegia)<sup>b</sup>** | 0.63 | 0.38 | 1.04 | 0.07 |
| **Diplegia (Ref: Quadriplegia)<sup>b</sup>** | 1.01 | 0.72 | 1.42 | 0.95 |
| **Triplegia (Ref: Quadriplegia)<sup>b</sup>** | 0.48 | 0.26 | 0.88 | **0.02** |
| **Gestational age: ≥37 weeks** | 4.68 | 3.52 | 6.24 | **0.00** |
| **Tone/Movement: Dystonia** | 1.02 | 0.77 | 1.35 | 0.91 |
| **Tone/Movement: Hypotonia** | 4.39 | 3.25 | 5.94 | **0.00** |
| **Tone/Movement: Other** | 2.16 | 1.40 | 3.33 | **0.00** |
| **Tone/Movement: Spasticity** | 0.68 | 0.48 | 0.96 | **0.03** |
| **GMFCS Level I (Ref: V)<sup>c</sup>** | 0.68 | 0.42 | 1.09 | 0.11 |
| **GMFCS Level II (Ref: V)<sup>c</sup>** | 1.21 | 0.80 | 1.82 | 0.37 |
| **GMFCS Level III (Ref: V)<sup>c</sup>** | 1.37 | 0.89 | 2.11 | 0.16 |
| **GMFCS Level IV (Ref: V)<sup>c</sup>** | 1.03 | 0.71 | 1.49 | 0.88 |

Individuals with genetic etiologies also had a significantly greater number of tone types documented than those without a genetic etiology (t-test p<0.001) (Table 5). Dystonia, hypotonia, and other tone/movement types (ataxia, chorea, or athetosis) typically presented in combination with each other or spasticity regardless of genetic etiology (Table 7).

**Table 7.** Percentage of individuals with each tone/movement type present in isolation (only) or in combination (mixed) stratified by presence vs. absence of a genetic etiology.

| Genetic etiology | Dystonia |  |  |  | Hypotonia |  |  |  | Other (Ataxia, Chorea, Athetosis) |  |  |  | Spasticity |  |  |  |
| --- | --- | --- | --- | --- | --- | --- | --- | --- | --- | --- | --- | --- | --- | --- | --- | --- |
|  | Only | % | Mixed | % | Only | % | Mixed | % | Only | % | Mixed | % | Only | % | Mixed | % |
| No | 33 | 4% | 893 | 96% | 170 | 28% | 440 | 72% | 19 | 14% | 118 | 86% | 1976 | 65% | 1057 | 35% |
| Yes | 1 | 1% | 151 | 99% | 77 | 31% | 171 | 69% | 8 | 13% | 52 | 87% | 99 | 35% | 186 | 65% |

However, spasticity was typically described in isolation when a genetic etiology was absent and typically described in combination with other tone/movement types when a genetic etiology was present (Table 7).

## Discussion

We have described the demographics and CP characteristics of 9,756 individuals with CP cared for across 22 tertiary care US medical sites using the CPRN clinical registry. To our knowledge, this is the largest population of individuals with CP in the US characterized using prospectively entered data. Logistic regression analysis of this data demonstrated that CP distribution, gestational age at birth, and tone/movement types are associated with GMFCS level and the presence of a genetic etiology. These data additionally show that Black race and age are associated with GMFCS level. These results add to our understanding of CP in the US, including by identifying race and age-based disparities in gross motor function and features associated with genetic CP etiologies.

### Race

In the 2020 US Census, the US population self-identified as 74.8% White, 13.7% Black, and 11.5% other races.^14^ The CPRN registry population assessed in this study were identified as 73.0% White, 18.7% Black, and 8.3% other races (Table 1). The higher percentage of Black individuals in the CPRN registry compared to the general US population may reflect the known higher prevalence of CP in Black individuals in the US, but warrants further study.^15^

Compared to White individuals with CP, our analysis showed that Black individuals with CP are twice as likely to have greater gross motor functional impairment (to be at GMFCS Level V vs. Levels I-II, Table 3). This may be associated with social drivers of health that correlate with race and may prevent equitable access to maternal care and/or CP therapies. Future work should determine whether race remains independently associated with gross motor impairment even when accounting for socio-economic status, which is separately associated with gross motor impairment in CP.^16^

### Age

The association between age and GMFCS level may be due to improved care over time for children with CP, barriers to ongoing care for adults with CP, and/or selection bias in our sample. Compared to adults, younger children may have had increased access to early therapeutic interventions known to reduce gross motor impairment in CP or may have been diagnosed with CP earlier despite milder impairments.^17,18^

Approximately one-third of adults with CP lose a GMFCS level as they age, a phenomenon attributed to early aging and loss of medical care and therapies for adults with CP.^19^ Adults represent 26.3% of the individuals in this study, even though the 22 contributing sites largely focus on pediatric care (Supplemental Table 1). Adults with greater gross motor impairments may be more likely to remain at pediatric sites for ongoing care. Overall, the high proportion of adults in this study support ongoing optimization of pediatric-to-adult care transition for individuals with CP.^20,21^

### Tone/movement types

Most individuals with CP in this study had spasticity (83.8%), matching the expected 85% reported in other population-based studies.^1,22–24^ There is less population-based data available, especially in the US, for the overall prevalence of other tone/movement types in CP (as opposed to their prevalence as the predominant tone type alone). Hypotonia (14.1%) and dystonia (31.8%) are common features of CP in this cohort while ataxia, chorea, and athetosis are less frequent (4.0%). Our data provide multi-site corroboration of single site observations demonstrating a high frequency of mixed tone/movement types.^25,26^ These data support considering all tone/movement types in an individual when describing CP characteristics or pursuing representative clinical trial recruitment.

### Associations with gross motor function

The CPRN registry matches the U-shaped distribution for GMFCS level noted in other population-based studies but demonstrates a higher proportion of individuals at GMFCS level V (23.5% in the CPRN registry vs. 13%–17% in other studies).^3,27–29^ This may be due to contributing sites being tertiary care medical sites and thus caring for individuals with greater medical complexity or due to inclusion of adults in this study, which may skew GMFCS to higher levels as described above.

We have shown that mixed tone/movement types (including spasticity, dystonia, and/or hypotonia) and birth at term gestation are more common in individuals with greater gross motor functional impairments (Tables 2 and 4). The presence of multiple tone/movement types may directly contribute to gross motor impairment or may be associated with more diffuse brain injury patterns that cause greater gross motor impairment. Dystonia is known to be more common in individuals with greater gross motor impairment,^30^ but this has been less well-established for hypotonia, as demonstrated here. Our data also demonstrate that the proportion of individuals with spasticity increases with higher GMFCS levels.

However, this increase is relatively small compared to the rise in proportions of individuals with dystonia and/or hypotonia with higher GMFCS levels (Table 2). We hypothesize that the modest increase in spasticity may be due to it being increasingly documented as an accompanying tone type alongside other tone/movement disorders at higher GMFCS levels (Table 4), but this warrants further study.

Compared to those born preterm, individuals with CP born at term gestation are almost twice as likely to be at GMFCS level V than at any other GMFCS level. This finding may be driven by an association between gestational age and CP etiology including the resultant brain injury patterns. For example, in other smaller studies, infection as a CP etiology and basal ganglia/thalamic injury on brain MRI independently predict the ability to walk, but gestational age does not.^26,31^ The association between GMFCS, CP etiology, and brain injury patterns warrant further investigation within the much larger CPRN registry.

### Associations with genetic etiology

Identification of a genetic etiology in 12% of individuals in the CPRN registry matches published estimates ranging from 8-30%.^26,32–34^ Notably, genetic etiology was identified by the treating clinician without standardization across individuals by genetic testing modality or access. With this critical caveat in mind, our data show that mixed tone/movement types (including ataxia, chorea, athetosis, and/or hypotonia) and birth at term gestation are associated with a clinician-reported genetic CP etiology (Tables 6 and 7). Based on the high yield of genetic testing and high rates of resultant changes in management (estimated at 8-10%) in individuals with CP, ^32,35^ recommendations are increasingly to pursue genetic testing for all individuals with CP.^35,36^ However, these recommendations outpace access to genetic testing and genetic counseling in many regions of the US.^37^ In situations where genetic testing access is limited, our results may help inform which individuals might be most likely to have a genetic etiology.

## Limitations

The CPRN clinical registry collects data from large tertiary care medical sites. Though these sites have large geographic catchment areas and are distributed across the US, they may not be representative of smaller community practices which may be more likely to provide CP care for individuals with less medical complexity or functional impairment. Geographical differences in CP demographics should be assessed in the future with greater outreach to community-based practices and the CP community itself.

Rates of data missingness varied across registry elements (Table 1). Though significant differences did exist between individuals with a complete data set (n=3057) and individuals with any data entered (n=9756), these differences were overall small and significance may have been driven by large sample sizes.^38^ Information on genetic etiology and gestational age was missing for almost half of individuals, but their proportions did not significantly differ between the individuals with a complete data set and the larger population (Table 1). Previous work utilizing the CPRN community registry has demonstrated that CP etiology is likely under-documented clinically,^39^ which would hamper data entry into a centralized clinical registry. Our analysis suggests the need for improved clinical documentation of CP etiology and gestational age at birth, which can prompt quality improvement initiatives across participating sites in a learning health network like CPRN.

Finally, any contemporary study characterizing CP must contend with evolving diagnostic approaches and CP descriptions.^40–42^ Learning health networks like CPRN are poised to rapidly implement changes to standardized CP diagnosis across its affiliated sites and examine any resultant changes in CP characteristics. As the clinical approach to CP diagnosis evolves, it is critical to establish CP characteristics as they present today to benchmark comparison to future cohorts.

## Conclusions

We report on the demographics and CP characteristics of a large cohort of individuals with CP and examine how these factors are associated with gross motor functional impairment and genetic etiology. This analysis is based on prospectively entered data from 22 tertiary care medical sites across the US and, to our knowledge, is the largest such study in the country. Our findings highlight racial and age-based disparities in gross motor function, suggest which individuals with CP may be most likely to have genetic etiologies, and establish benchmarks for advancing CP care in the US.

## Data Availability

All data produced in the present study are available upon reasonable request to the Cerebral Palsy Research Network Data Coordinating Center via the authors.

## Acknowledgments

We are grateful for the following individuals who contributed to data collection at their CPRN sites: Jim McCarthy, Jeffrey Leonard, Tom Novachek, Cynthia Wozow, Shenandoah Robinson, Eric Chin, and Robert Bollo.

## Abbreviations

CP: cerebral palsy
CPRN: Cerebral Palsy Research Network
GMFCS: Gross Motor Function Classification System

---

**Supplementary Table 1.**
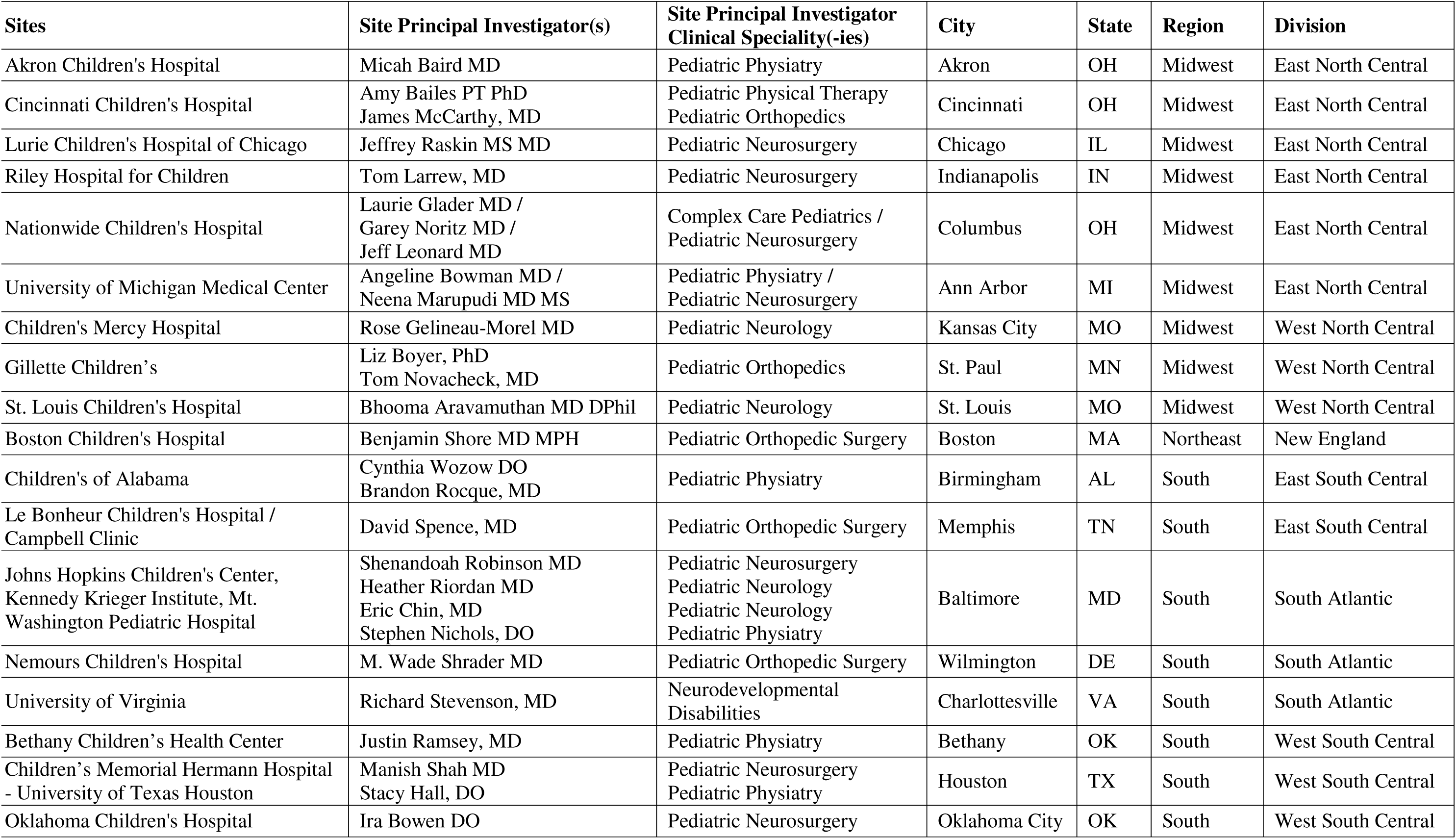
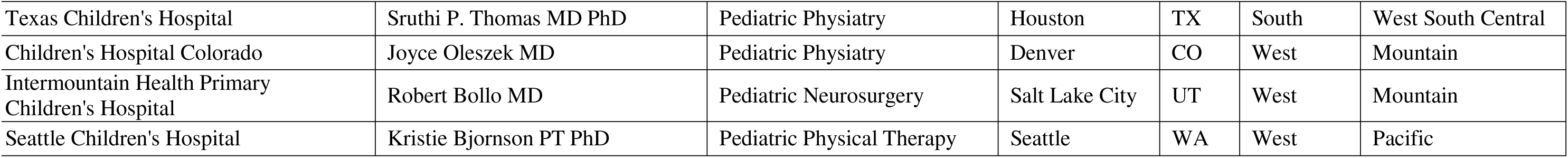
Cerebral Palsy Research Network study sites. Regions and Divisions are per US Census designations.

## Notes

### Competing Interest Statement

The authors have declared no competing interest.

### Funding Statement

Pediatric Epilepsy Research Foundation (BRA), St. Louis Children's Hospital Foundation (BRA)

### Author Declarations

The Institutional Review Board (IRB) of Nationwide Children's Hospital gave ethical approval for this work.

